# Dynamics of immune recall following SARS-CoV-2 vaccination or breakthrough infection

**DOI:** 10.1101/2021.12.23.21268285

**Authors:** Marios Koutsakos, Wen Shi Lee, Arnold Reynaldi, Hyon-Xhi Tan, Grace Gare, Paul Kinsella, Kwee Chin Liew, Deborah A. Williamson, Helen E. Kent, Eva Stadler, Deborah Cromer, David S. Khoury, Adam K. Wheatley, Jennifer A. Juno, Miles P. Davenport, Stephen J. Kent

**Affiliations:** Department of Microbiology and Immunology, University of Melbourne, Peter Doherty Institute for Infection and Immunity, Melbourne, Victoria, Australia; Kirby Institute, University of New South Wales, Kensington, New South Wales, Australia; Victorian Infectious Diseases Reference Laboratory, The Royal Melbourne Hospital at The Peter Doherty Institute for Infection and Immunity, Melbourne, VIC, Australia; Department of Infectious Diseases, The University of Melbourne at the Peter Doherty Institute for Infection and Immunity, Victoria, 3000, Australia; Melbourne Sexual Health Centre and Department of Infectious Diseases, Alfred Hospital and Central Clinical School, Monash University, Melbourne, Victoria, Australia

## Abstract

Vaccination against SARS-CoV-2 results in protection from acquisition of infection as well as improved clinical outcomes even if infection occurs, likely reflecting a combination of residual vaccine-elicited immunity and the recall of immunological memory. Here, we define the early kinetics of spike-specific humoral and T cell immunity after vaccination of seropositive individuals, and after breakthrough infection in vaccinated individuals. Intensive and early longitudinal sampling reveals the timing and magnitude of recall, with the phenotypic activation of B cells preceding an increase in neutralizing antibody titres. In breakthrough infections, the delayed kinetics of humoral immune recall provides a mechanism for the lack of early control of viral replication but likely underpins accelerated viral clearance and the protective effects of vaccination against severe COVID-19.

---

Vaccines encoding the spike (S) antigen of SARS-CoV-2 are effective in reducing the risk of symptomatic SARS-CoV-2 infection, as well as progression to severe COVID-19 disease (*1*). Neutralizing antibodies are a correlate of protection (*2, 3*) and likely act to prevent infection by blocking viral attachment and entry. However, as antibody levels naturally wane (*4*), vaccine effectiveness drops (*5*) and the frequency of “breakthrough infections” among vaccinated individuals increases in the population. The emergence of antigenic variants including Beta and Omicron have highlighted the potential for viral escape from neutralizing antibody recognition, which can considerably reduce vaccine effectiveness against acquisition of SARS-CoV-2 infection (*6*). Nevertheless, vaccine-elicited immunity continues to provide robust protection against severe disease outcomes, even in the face of viral variants (*7*). Viral growth rates and peak viral RNA levels in the upper respiratory tract are similar between vaccinated and unvaccinated infected individuals during the first week of infection (*8-10*). However, vaccinated individuals consistently display more rapid clearance of viral RNA than unvaccinated controls during the second week of infection (*8, 9*). Importantly, there is a lower probability of culturing infectious virus from respiratory samples of infected vaccinated individuals (*11*). However, the immunological mechanisms that underpin accelerated viral clearance remain unclear. The comparable viral levels within vaccinated and unvaccinated individuals in the first week of infection suggest that residual (post-vaccination, pre-infection) antibody or T cell immunity fails to limit early viral replication in the respiratory tract. However, the recall of SARS-CoV-2 specific antibodies, memory B and T cell responses following breakthrough infection could contribute to viral clearance and temper disease severity, as is thought to be the case for other respiratory viral infections (*12, 13*). Understanding the mechanisms and effectiveness of recall responses in protecting from severe SARS-CoV-2 infection is critical to informing the optimal deployment of current vaccines and guiding the design of novel vaccines to maintain maximal protection against severe disease. To date, however, the precise kinetics of immune recall in the context of SARS-CoV-2 infection are poorly resolved.

To understand the dynamics of recall of SARS-CoV-2 spike-specific immunity, we first analysed immune responses after vaccination of seropositive individuals. We recruited and longitudinally sampled a cohort of 25 individuals with previous PCR-confirmed SARS-CoV-2 infection and/or baseline spike protein seropositivity (seropositive group), and a comparator group of 8 seronegative individuals with no history of SARS-CoV-2 infection (naïve group) (Table S1). We undertook early longitudinal sampling from day 3 onwards after vaccination with either BNT162b2 or ChAdOx1 nCoV-19 vaccines. In seropositive individuals, S- and RBD-specific antibody titres began to increase 5 days after vaccination, with titres peaking between day 10-14 (Fig 1A and B). In naïve individuals, S and RBD antibody titres emerged later after the first dose (day 9 onwards) and remained at lower levels compared to immunized seropositive individuals, in line with other reports of primary immunization of immunologically naïve individuals (*14*). We also assessed serological responses using a live virus neutralization assay (*15*). At the time of vaccination, only 47% of previously infected individuals had detectable plasma neutralization activity, which reflected residual activity following waning from peak neutralization titres seen in early convalescence (Fig 1C). Following the first vaccine dose, neutralizing titres increased from day 6, concomitant with the rise in S and RBD binding antibodies and peaked between day 10-14 (Fig 1C). In contrast, immunization of naïve individuals elicited much lower levels of neutralizing antibodies, which only emerged around day 12 post-first dose. We applied a segmented linear model to estimate the initial period of delay, the rate of increase, and fold change over baseline in the recall of antibodies (Table S2). We estimated that the initial delay phase before neutralizing antibody levels increased was 4.85 days, with a doubling time of 0.74 days thereafter and with a peak fold-change over baseline of 25.8.

**Figure 1.**
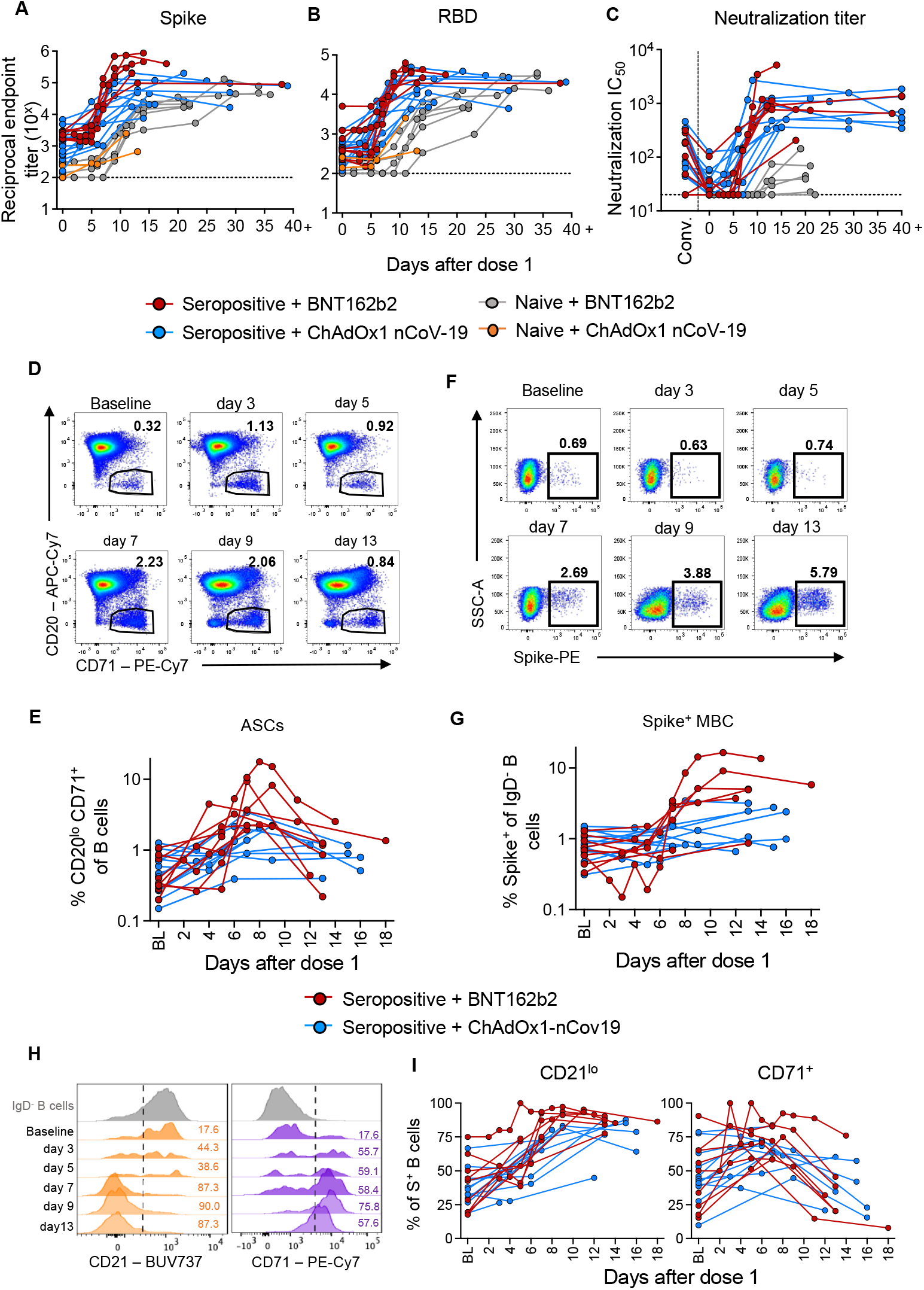
Recall kinetics of humoral immune responses following vaccination. (A-C) Serological analysis of samples following one dose of BNT162b2 or ChAdOx1 nCoV-19. Kinetics of S-specific IgG (A) and RBD-specific IgG (B) antibodies measured by ELISA and neutralizing antibodies measured by a live virus neutralization assay (C) in SARS-CoV-2 naïve or seropositive individuals (seropositive n=19, naïve n=8). (D-E) Representative flow cytometry plots (D) and frequency (E) of antibody-secreting cells (ASCs, CD20^lo^CD71^+^ B cells) in SARS-CoV-2 seropositive individuals. (F-G) Representative flow cytometry plots (F) and frequency (G) of S-specific class-switched B cells (IgD^-^CD19^+^) in vaccinated seropositive individuals. (H-I) Representative flow cytometry plots (H) and frequency (I) of activation markers (CD21, CD71) within S-specific class-switched B cells (IgD^-^CD19^+^) in vaccinated seropositive individuals. (D-I) n=21.

Memory B cells constitute an important arm of durable vaccine-elicited immunity, rapidly responding to secondary antigen exposure via differentiation into antibody secreting cells (ASCs). To better understand memory B cell re-activation in vivo, we assessed changes in the frequency and phenotype of SARS-CoV-2 specific memory B cells in seropositive individuals (n=21) in response to immunization. Antibody-secreting cells (ASCs; CD19^+^CD20^lo^CD71^+^, commonly termed plasmablasts) expanded in peripheral blood, increasing from as early as day 3 based on flow cytometry (estimated by segmented linear modelling to occur as early as day 2.4), peaking between day 7 and 9 before contracting to near baseline levels from day 11 onwards (Fig 1D and E). S-specific class-switched memory B cells (Spike^+^IgD^-^ CD19^+^ cells), which unlike ASCs constitute a stable population of quiescent memory (*4*), were detectable in all seropositive individuals prior to vaccination (0.31-1.5% of IgD^-^ B cells). Following vaccination, the frequency of S-specific class-switched B cells increased from day 7 onwards based on flow cytometry (estimated as early as day 6.5) and peaked by day 10 (Fig 1 F and G). The activation state of S-specific B cells was assessed longitudinally using surface-expressed activation markers CD21 and CD71 (*16*). Consistent with expansion of S-specific B cells, CD21 downregulation and CD71 upregulation, both denoting cellular activation, were evident as early as day 3 and was maximal around day 9 (Fig 1H-I).

Given the potential of T cells to contribute to the control of viral replication and the association of CD4^+^ T cell responses with the development of neutralizing antibodies (*17*), we assessed the recall of S-specific CD4^+^ and CD8^+^ T cells following vaccination of seropositive individuals. Using re-stimulation with recombinant S protein and an activation-induced marker (AIM) assay (Fig S1 and S2), an increase in both S-specific CD4^+^ memory T cells (CD4^+^Tmem; CD45RA^-^CXCR5^-^)(Fig 2A and B) and circulating CD4^+^ T follicular helper cells (cTFH; CD45RA^-^CXCR5^+^) was evident from day 5 onwards, peaking around day 9 and declining thereafter (Fig 2C and D). Recall of S-specific CD4^+^ T cell responses have also been previously quantified at an epitope-specific level in a subset of the vaccination cohort (n=10 individuals; (*18*)). Use of an HLA-DRB1*15/S_751_ tetramer to precisely enumerate antigen-specific T cell frequencies following vaccination provided similar results to the AIM assay, with recall evident from day 5 onward and peaking between days 8 and 10 (Fig 2E and F). Similar kinetics were observed for S-specific CD8^+^ memory T cells (Fig 2G and H), albeit at a lower magnitude than S-specific CD4^+^ T cell responses. Using the same modelling approach as above, the initial delay for T cell recall was ∼4 days (Table S2). The peak levels (amongst available samples) of ASCs and S-specific cTFH cells were positively correlated with the peak binding and neutralizing antibodies, as well as with each other (Fig S3), consistent with data from primary infection and vaccination (*17*).

**Figure 2.**
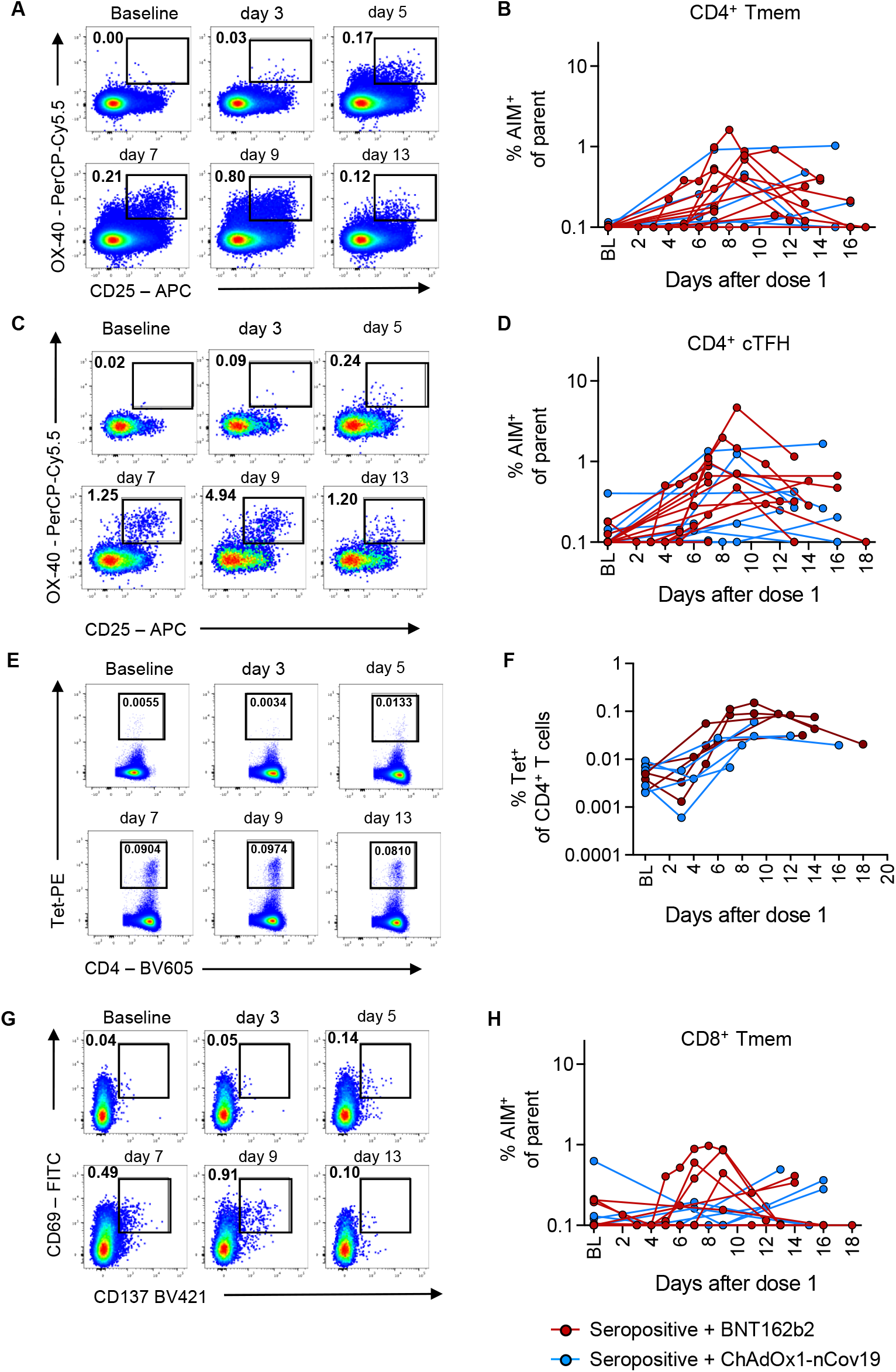
Recall kinetics of memory T cell activation following vaccination. Analysis of S-specific T cells by AIM assay following one dose of BNT162b2 or ChAdOx1 nCoV-19 in SARS-CoV-2 naïve or seropositive individuals. (A, C, E, G) Representative staining and frequency of AIM markers (CD25, OX-40) on CD4^+^ Tmem cells (CD3^+^CD4^+^CD8^-^CD45RA^-^CXCR5^-^) (A-B) and CD4^+^ cTFH cells (CD3^+^CD4^+^CD8^-^CD45RA^-^CXCR5^+^) (C-D), tetramer staining (E-F) or AIM markers (CD69, CD137) on CD8^+^ Tmem cells (CD3^+^CD8^+^CD4^-^non-naüve) (G-H) after stimulation with 5μg/ml of SARS-CoV-2 S protein on different timepoints after vaccination.

While vaccination of previously infected individuals provides a tractable model to assess immunological recall, the extent to which it recapitulates the dynamics of actual breakthrough infection of vaccinated individuals is unclear. Studies of the early immune kinetics of breakthrough infection are challenging, as the timing of initial infection is rarely known and is often referenced from the time of symptom onset (estimated to be a mean of 4.3 days after acquisition for Delta) (*19*). Nevertheless, we recruited 6 individuals with generally mild-moderate PCR-confirmed breakthrough SARS-CoV-2 infections that occurred 1-4 months after receiving a second dose of a COVID vaccine (Table S3), during a time when Delta was the dominant circulating variant. Serial blood samples and nose swabs were obtained over 1-18 days after symptom onset and S-specific antibody and cellular immune responses analysed as before. An additional follow-up sample was available for four donors between day 26-39. For four individuals, the time of infection could be definitively established as 2-3 days prior to symptom onset as these subjects had a single defined exposure event in a low incidence environment. Neutralizing antibody titres, as well as S- and RBD-binding antibodies, remained at baseline levels for a remarkable 5-7 days after symptom onset (8-10 days post exposure) before rising steadily during the second and third weeks up to the last time point collected (Fig 3A-C). Activation of S-specific memory B cells was evident from day 6 post symptom onset (Fig S3A). The circulating frequencies of ASCs and S-specific memory B cells remained stable for the first week after symptom onset, before expanding around day 7-8 post symptom onset (Fig 3C and D). With the exception of subjects #5 and #6, whose precise exposure time was undefined, the recall kinetics of antibody and memory B cells following breakthrough infection appeared delayed when compared to vaccination of seropositive individuals. Surprisingly, frequencies of S-specific CD4^+^ and CD8^+^ T cells remained largely unchanged following breakthrough infection in 5 of 6 individuals, with only a single subject (#6) displaying a >5-fold increase (Fig 3E-G). These results are in stark contrast to the rapid and clear recall of CD4^+^ and CD8^+^ T cell immunity following vaccination of seropositive subjects shown in Fig 2.

**Figure 3.**
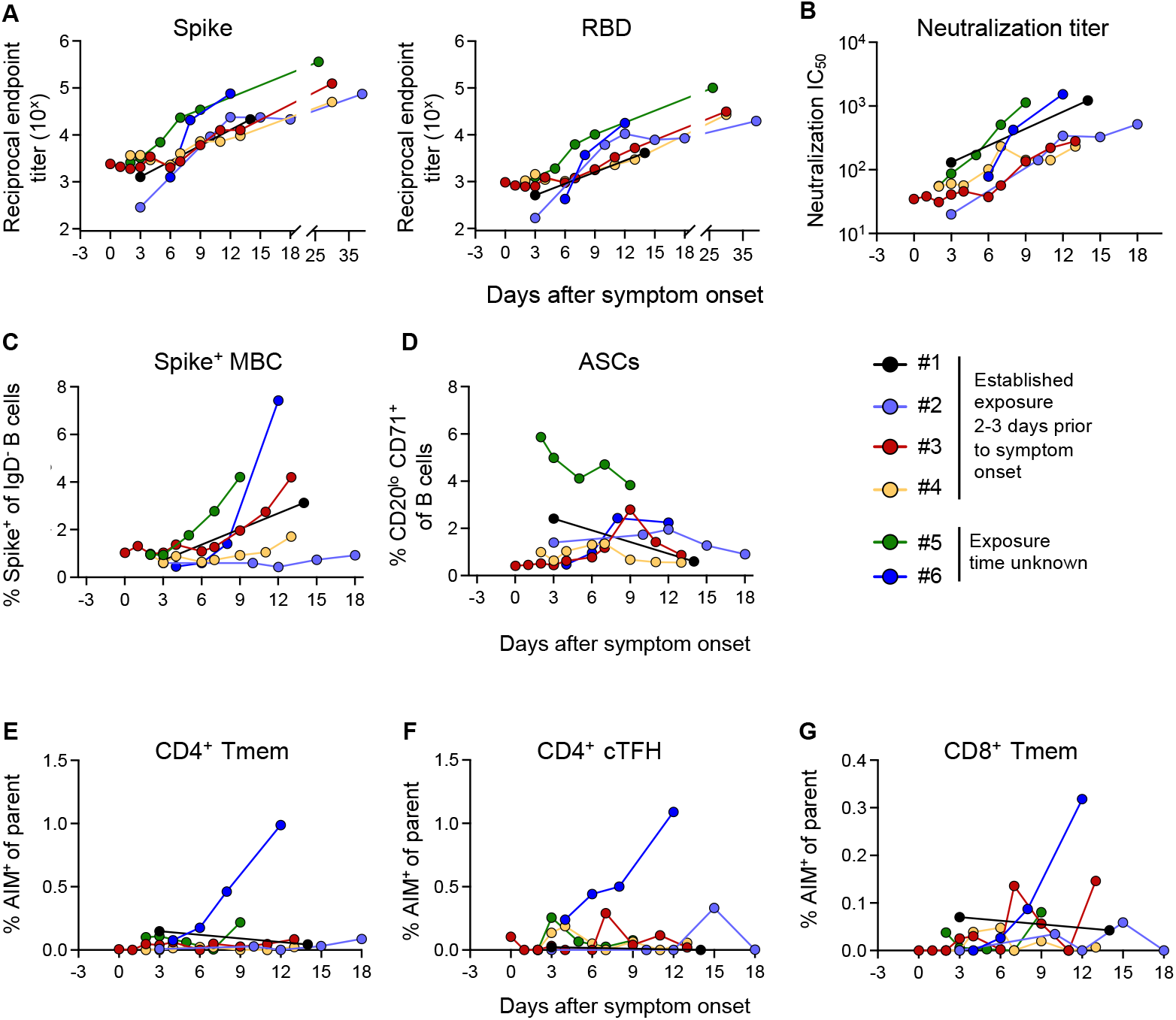
Recall kinetics in SARS-CoV-2 breakthrough infections. (A) Kinetics of S- and RBD-specific IgG antibodies measured by ELISA and (B) of neutralizing antibodies measured by a live virus microneutralization assay. (C) Frequencies of S-specific class-switched B cells (IgD^-^CD19^+^) and (D) antibody-secreting cells (ASCs, CD20^lo^CD71^+^ B cells) determined by flow cytometry. (E) Frequencies of AIM^+^ (CD25, OX-40) S-specific CD4^+^ Tmem cells (CD3^+^CD4^+^CD8^-^CD45RA^-^CXCR5^-^), (F) CD4^+^ cTFH cells (CD3^+^CD4^+^CD8^-^CD45RA^-^CXCR5^+^) and (G) AIM^+^ (CD69, CD137) S-specific CD8^+^ Tmem cells (CD3^+^CD8^+^CD4^-^non-naüve) after stimulation with 5μg/ml of SARS-CoV-2 S protein. N=6 participants. The x-axis is set as days post symptom onset with negative values indicating the confirmed exposure time for 4 participants (2-3 days prior to symptom onset).

To compare the dynamics of immune recall following vaccination and breakthrough infection, we applied the same model to parameterize the kinetics of recall. We considered the delay from exposure to symptom onset (which was known in 4/6 subjects and conservatively assumed this to also be 3 days prior to symptom onset in the other 2 subjects). The estimated time to initial increase in neutralizing and binding antibody levels was 7.22 and 8.21 days after exposure respectively, and 9.3 days for S-specific B cells (Table S4), all of which were delayed compared to vaccination (increase estimated prior to day 7 for all three measures, Fig 4A).

**Figure 4.**
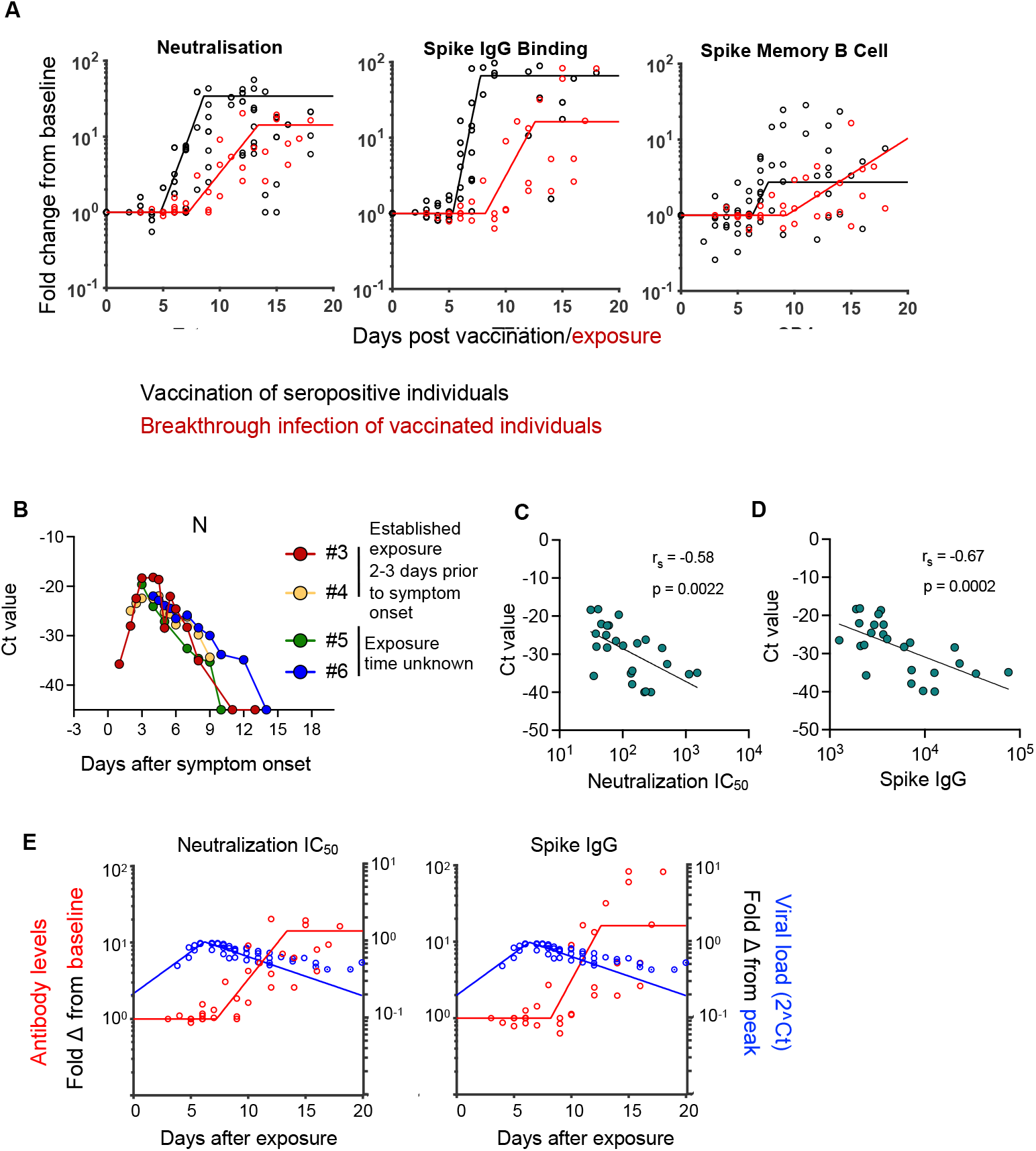
Immune recall and viral load kinetics in SARS-CoV-2 breakthrough infections. (A) Comparative kinetics of immune recall following vaccination of seropositive individuals (black) and breakthrough infection of vaccinated individuals (red). (B) Ct values for SARS-CoV-2 N gene in serial nasopharyngeal swabs. (C-D) Correlations between Ct value and levels of neutralizing antibody titres (C) or spike IgG antibodies (D). Spearman correlation coefficients (r_s_) and p values are indicated on the figure, n=25 paired samples from 4 subjects. (E) Overlayed kinetics of antibody recall (fold change over baseline, in red) and viral load (fold change relative to peak, in blue) for neutralizing antibody titres and spike IgG antibodies.

To investigate the relationship between recall immunity and viral control, we analysed viral load kinetics by qPCR of N (Fig 4B), RDRP and S (Fig S4B-E) genes in serial nasopharyngeal swabs from 4 of the individuals with breakthrough infection. This indicated a peak of viral replication (amongst available timepoints) on day 7-8 after infection (day 4-5 after symptom onset), followed by rapid viral clearance thereafter. Neutralizing and binding antibody titres were negatively correlated with viral loads (Fig 4C and D, Fig S4C-G). Comparison of viral load kinetics with the recall of antibodies, indicated that the peak of viral load preceded the rise in neutralizing antibodies and that recall of humoral immunity coincided with a decrease in viral load (Fig 4E).

Overall, intensive longitudinal sampling during vaccination of SARS-CoV-2 seropositive subjects and breakthrough infection revealed the sequence and dynamics of recalled immune memory to SARS-CoV-2 S protein. Following vaccination, phenotypic activation of S-specific memory B cells coincides with the rapid expansion of ASCs (as early as day 3 after vaccination) and is followed by an increase of antigen-specific T cells in the blood (day 5 onwards). Subsequently, serological titres of both binding and neutralizing antibodies rapidly increase (day 5 onwards) and are stably maintained for at least 30 days after antigen re-exposure, with increases in frequencies of antigen-specific memory B cells following a similar trajectory. Although recall dynamics of antibody responses were relatively uniform in the context of vaccination, during breakthrough infection of vaccinated individuals this was more variable in timing and magnitude. This likely reflects the inherent heterogeneity of viral replication and timing of symptom onset between individuals. Where the timing of exposure was defined, the delay between infection and antibody recall was ∼7-8 days (including 2-3 prior and 5-6 days after symptom onset). The longer delay in recall of immunity following infection compared to vaccination may reflect differences in antigen accumulation between respiratory acquisition of infection compared to the bolus introduction of S-encoding mRNA or adenovirus following intramuscular vaccination. Nonetheless, the kinetics of recall of immune memory in SARS-CoV-2 is broadly consistent with studies of human influenza infection (*20, 21*).

Previous studies show viral growth rates and peak viral loads are comparable between vaccinated and unvaccinated individuals following SARS-CoV-2 infection, although vaccinated subjects show a faster viral clearance after the peak (*8, 9*). A major question is to identify which immune responses mediate rapid viral clearance and if this is mechanistically linked to protection from severe infection in vaccinated subjects. Several lines of evidence suggest neutralizing antibody responses may play a major role in protection from severe disease. For example, early treatment of COVID-19 patients with potently neutralizing monoclonal antibody treatments significantly reduces the risk of progression to severe disease (*22, 23*). Protection is generally not observed following analogous administration of convalescent plasma, suggesting while antibodies alone appear sufficient to moderate disease severity, neutralization potency is likely a critical determinant (*24, 25*). Despite the non-trivial differences between recall of endogenous antibody and passive antibody therapy, the timing and magnitude of the increase in neutralizing antibody levels in breakthrough infection constitute a plausible modality for the reduced risk of severe disease observed in population studies of breakthrough infections (*26*). Although systemic recall of T cell responses was not detected in the first 18 days in 5 of 6 of breakthrough infections studied, we cannot preclude T cell migration to the site of infection. Therefore, any contribution of T cells in mitigating disease severity of breakthrough infections requires investigation in larger cohorts, with particular attention paid to SARS-CoV-2 specific T cells in the respiratory tract which would be favourably localized to temper viral replication.

An important caveat of our study is the lack of mucosal sampling of the upper or lower respiratory tracts. Studies of intranasal SARS-CoV-2 challenge after intramuscular vaccination of non-human primates (*27, 28*) indicate a temporal disconnect in recall kinetics between anatomical compartments, with both SARS-CoV-2 specific and non-specific IgG levels increasing in the bronchoalveolar lavage fluid (although this may reflect inflammation-induced exudation) prior to antibody recall in the serum. Additionally, the accelerated viral clearance observed in vaccinated individuals (*8, 9*) is based on analysis of nasopharyngeal samples, which may not necessarily reflect the viral loads or kinetics of the lower respiratory tract.

Encouragingly, we find breakthrough infection of vaccinated individuals drives re-expansion of humoral immune memory with augmented neutralizing antibodies, albeit with some delay. This suggests that recall of immunity may mitigate disease severity of breakthrough infections with antigenically distinct variants including Omicron, while also boosting population level immunity against SARS-CoV-2, potentially further restricting the healthcare burden inflicted by the pandemic and smoothing a pathway towards endemicity.

## Data Availability

All data produced in the present study are available upon reasonable request to the authors

## Acknowledgements

We thank the participants for the generous involvement and provision of samples. We thank T. Amarasena, R. Esterbauer, K. Wragg, P. Konstandopoulos, K. Field and A. Kelly (University of Melbourne) for excellent technical assistance. We thank molecular staff at the Victorian Infectious Diseases Reference Laboratory for performing RT-PCR. We acknowledge the Melbourne Cytometry Platform (Melbourne Brain Centre node) for provision of flow cytometry services.

## Author contributions

Conceptualization: MK, WSL, AKW, JAJ, MPD, SJK

Formal Analysis: MK, WSL, AR, KCL, ES, DC, DSK, AKW, JAJ

Funding acquisition: AKW, JAJ, MPD, SJK

Investigation: MK, WSL, AR, HXT, GG, KCL, AKW, JAJ,

Methodology: MK, WSL, AR, HXT, GG, KCL, AKW, JAJ

Resources: PK, KCL, DAW, HEK, SJK

Supervision: DC, DSK, AKW, JAJ, MPD, SJK

Writing – original draft: MK, WSL, AKW, JAJ, MPD, SJK

Writing – review & editing: MK, WSL, AR, HXT, DAW, DC, DSK, AKW, JAJ, MPD, SJK

## Data and materials availability

All data are available in the main text or the supplementary materials.

## Supplementary Materials

### Materials and Methods

#### Ethics Statement

The study protocols were approved by the University of Melbourne Human Research Ethics Committee (2021-21198-15398-3, 2056689), and all associated procedures were carried out in accordance with approved guidelines. All participants provided written informed consent in accordance with the Declaration of Helsinki.

#### Participant recruitment and sample collection

A cohort of subjects with either a prior positive nasal PCR for SARS-CoV-2 or a positive ELISA for SARS-CoV-2 S and RBD protein were recruited to provide blood samples following vaccination against SARS-CoV-2. Contemporaneous controls who had not previously experienced any symptoms of COVID-19 and who were confirmed to be seronegative were also recruited to provide blood samples prior to and following vaccination for SARS-CoV-2 (Table S1). A cohort of previously vaccinated participants with a nasal PCR-confirmed breakthrough COVID-19 were recruited through contacts with the investigators and invited to provide serial blood samples (Table S1). For all participants, whole blood was collected with sodium heparin anticoagulant. Plasma was collected and stored at −80 °C, and PBMCs were isolated via Ficoll-Paque separation, cryopreserved in 10% DMSO/FCS and stored in liquid nitrogen.

#### ELISA

Antibody binding to SARS-CoV-2 S or RBD proteins was tested by ELISA. The expression of recombinant S and RBD has been described previously (*29*). For ELISA, 96-well Maxisorp plates (Thermo Fisher) were coated overnight at 4°C with 2 μg/ml recombinant S or RBD proteins. After blocking with 1% FCS in phosphate-buffered saline (PBS), duplicate wells of serially diluted plasma were added and incubated for 2 h at room temperature. Plates were washed in PBS-T (0.05% Tween-20 in PBS) and PBS before incubation with 1:20,000 dilution of HRP-conjugated anti-human IgG (Sigma) for 1 h at room temperature. Plates were washed and developed using TMB substrate (Sigma), stopped using sulphuric acid and read at 450 nm. Endpoint titers were calculated as the reciprocal serum dilution giving signal 2× background using a fitted curve (4 parameter log regression).

#### Microneutralization assay with ELISA-based read out

Plasma neutralization activity against SARS-CoV-2 was measured using a microneutralization assay as previously described (*15*). Wildtype SARS-CoV-2 (CoV/Australia/VIC/01/2020) isolate was passaged in Vero cells and stored at -80ºC. 96-well flat bottom plates were seeded with Vero cells (20,000 cells per well in 100µl). The next day, Vero cells were washed once with 200 µl serum-free DMEM and added with 150µl of infection media (serum-free DMEM with 1.33 µg/ml TPCK trypsin). 2.5-fold serial dilutions of heat-inactivated plasma (1:20-1:12207) were incubated with SARS-CoV-2 virus at 2000 TCID_50_/ml at 37ºC for 1 hour. Next, plasma-virus mixtures (50µl) were added to Vero cells in duplicate and incubated at 37°C for 48 hours. ‘Cells only’ and ‘virus+cells’ controls were included to represent 0% and 100% infectivity respectively. After 48 hours, all cell culture media were carefully removed from wells and 200 µl of 4% formaldehyde was added to fix the cells for 30 mins at room temperature. The plates were then dunked in a 1% formaldehyde bath for 30 minutes to inactivate any residual virus prior to removal from the BSL3 facility. Cells were washed once in PBS and then permeabilised with 150µl of 0.1% Triton-X for 15 minutes. Following one wash in PBS, wells were blocked with 200µl of blocking solution (4% BSA with 0.1% Tween-20) for 1 hour. After three washes in PBST (PBS with 0.05% Tween-20), wells were incubated with 100µl of rabbit polyclonal anti-SARS-CoV N antibody (Rockland, #200-401-A50) at a 1:8000 dilution in dilution buffer (PBS with 0.2% Tween-20, 0.1% BSA and 0.5% NP-40) for 1 hour. Plates were then washed six times in PBST and added with 100µl of goat anti-rabbit IgG (Abcam, #ab6721) at a 1:8000 dilution for 1 hour. After six washes in PBST, plates were developed with TMB and stopped with 0.15M H_2_SO_4_. OD values read at 450nm were then used to calculate %neutralization with the following formula: (‘Virus + cells’ – ‘sample’) ÷ (‘Virus + cells’ – ‘Cells only’) × 100. IC_50_ values were determined using four-parameter nonlinear regression in GraphPad Prism with curve fits constrained to have a minimum of 0% and maximum of 100% neutralization.

#### Flow cytometric detection of SARS-CoV-2-reactive B cells

Probes for delineating SARS-CoV-2 S-specific B cells within cryopreserved human PBMCs were generated by sequential addition of streptavidin-phycoerythrin (PE) (Thermo Fisher) to trimeric S protein biotinylated using recombinant Bir-A (Avidity). Cells were stained with Aqua viability dye (Thermo Fisher) in PBS. Cells were then stained with S-PE probes and surface monoclonal antibodies in 1% FCS in PBS for 30 mins at 4°C. Monoclonal antibodies for surface staining included CD19-ECD (J3-119, 1:150) (Beckman Coulter), IgM BUV395 (G20-127, 1:150), CD21 BUV737 (B-ly4, 1:150), IgG BV786 (G18-145, 1:75), streptavidin-BV510 (1:600), CD11c (B-ly6, 1:100) (BD Biosciences), CD20 APC-Cy7 (2H7, 1:150), CD14 BV510 (M5E2, 1:300), CD3 BV510 (OKT3, 1:600), CD8a BV510 (RPA-T8, 1:1500), CD16 BV510 (3G8, 1:500), CD10 BV510 (HI10a, 1:750) and CD27 BV605 (O323, 1:150), CD71 PeCy7 (CY1G4, 1:100) (BioLegend), IgD AF488 (Goat polyclonal, 1:100) (Southern Biotech), IgA VioBlue (IS11-8E10, 1:100) (Miltenyi Biotec). Cells were washed, fixed with 1% formaldehyde (Polysciences) and acquired on a BD LSR Fortessa.

#### Flow cytometric detection of antigen-specific T cells

Analysis of SARS-CoV-2 specific T cells was performed as previously described (*29*). Briefly, cryopreserved human PBMCs were thawed and rested for 4 h at 37°C. Cells were cultured in 96-well plates at 0.5-4 × 10^6^ cells per well and stimulated for 20 h with 5 μg ml^−^1 of protein (BSA, SARS-CoV-2 S). Cells from selected donors were also stimulated with SEB (5 μg ml^−^1) as a positive control. An CD154 APC-Cy7 (TRAP-1, BD Biosciences) antibody was included in the culture medium for the duration of the stimulation. After stimulation, cells were washed, stained with Live/Dead blue viability dye (Thermo Fisher) and incubated in a cocktail of monoclonal antibodies: CD27 BV510 (L128, 1:50), CCR7 Alexa700 (150503, 1:50), CD45RA PE-Cy7 (HI100, 1:200), (BD Biosciences), CD3 BUV395 (SK7, 1:100), CD4 BV605 (RPA-T4, 1:100), CD8 BV650 (RPA-T8, 1:400), CD25 APC (BC96, 1:50), OX-40 PerCP-Cy5.5 (ACT35, 1:50), CCR6 BV785 (G034E3, 1:100), CXCR3 PE-Dazzle 594 (G02H57, 1:50), CD69 FITC (FN50, 1:200), CD137 BV421 (4B4-1, 1:100) (BioLegend) and CXCR5 PE (MU5UBEE, Thermo Fisher, 1:50). Cells were washed, fixed with 1% formaldehyde and acquired on a BD LSR Fortessa using BD FACS Diva. The HLA-DRB1*15/S751 tetramer (ProImmune) and associated data are described in Wragg et al (*18*). Briefly, cells were incubated with 50nM dasatinib for 30 minutes at 37ºC, then stained with PE-conjugated tetramer at 4ug/mL for 60 minutes at 37ºC. Cells were washed in PBS, labelled with Live/Dead green viability dye, and stained with a cocktail of surface antibodies for 30 minutes at 4ºC. Surface stain antibodies included: CD45RA PerCP-Cy5.5 (HI100), CD4 BV605 (RPA-T4), CD3 BUV395 (SK7) and CD20 BUV805 (2H7) (BD Biosciences).

### Analysis of viral RNA load by qPCR

#### Nucleic acid extraction and complementary DNA (cDNA) synthesis

For viral RNA extraction, 200 mL of sample was extracted with the QIAamp 96 Virus QIAcube HT kit (Qiagen, Germany) on the QIAcube HT System (Qiagen) according to manufacturer’s instructions. Purified nucleic acid was then immediately converted to cDNA by reverse transcription with random hexamers using the SensiFAST cDNA Synthesis Kit (Bioline Reagents, UK) according to manufacturer’s instructions. cDNA was used immediately in the rRT-PCR or stored at -20ºC.

#### SARS-CoV-2 rRT-PCR

Three microlitres of cDNA was added to a commercial real-time PCR master mix (PrecisionFast qPCR Master Mix; Primer Design, UK) in a 20 mL reaction mix containing primers and probe with a final concentration of 0.9 mM and 0.2 mM for each primer and the probe, respectively. Samples were tested for the presence of SARS-CoV-2 RNA-dependent RNA polymerase (RdRp)/helicase (Hel), spike (S), and nucleocapsid (N) genes using previously described primers and probes (*30, 31*).

Thermal cycling and rRT-PCR analyses for all assays were performed on the ABI 7500 FAST real-time PCR system (Applied Biosystems, USA) with the following thermal cycling profile: 95C for 2 min, followed by 45 PCR cycles of 95C for 5 s and 60C for 25 s for N gene and 95C for 2 min, followed by 45 PCR cycles of 95C for 5 s and 55C for 25 s for RdRP/Helicase gene and S gene.

### Modelling the kinetics of immune recall

We used a segmented model to estimate the activation time and growth rate of various immune responses after vaccination and breakthrough infection. The model of the immune response *y* for subject *i* at time *y*_*i*_ can be written as:

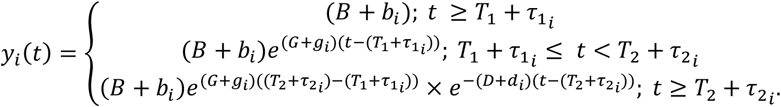

The model has 5 parameters; *B, G, T*_1_, *D*, and *T*_2_. For a period before *T*_1_, we assumed a constant baseline value *B* for the immune response. After the activation time *T*_1_, the immune response will grow at a rate of *G* until *T*_2_. From *T*_2_, the immune response will decay at a rate of *D*. For each subject *i*, the parameters were taken from a normal distribution, with each parameter having its own mean (fixed effect). A diagonal random effect structure was used, where we assumed there was no correlation within the random effects. The model was fitted to the log-transformed data values, with a constant error model distributed around zero with a standard deviation *σ*. To account for the values less than the limit of detection, a censored mixed effect regression was used to fit the model. Values less 20, 100, and 0.0001 were censored for the neutralization, IgG bindings, and T cell data respectively. Model fitting was performed using MonolixR2019b.

## Supplementary Figures

**Supplementary Figure 1.**
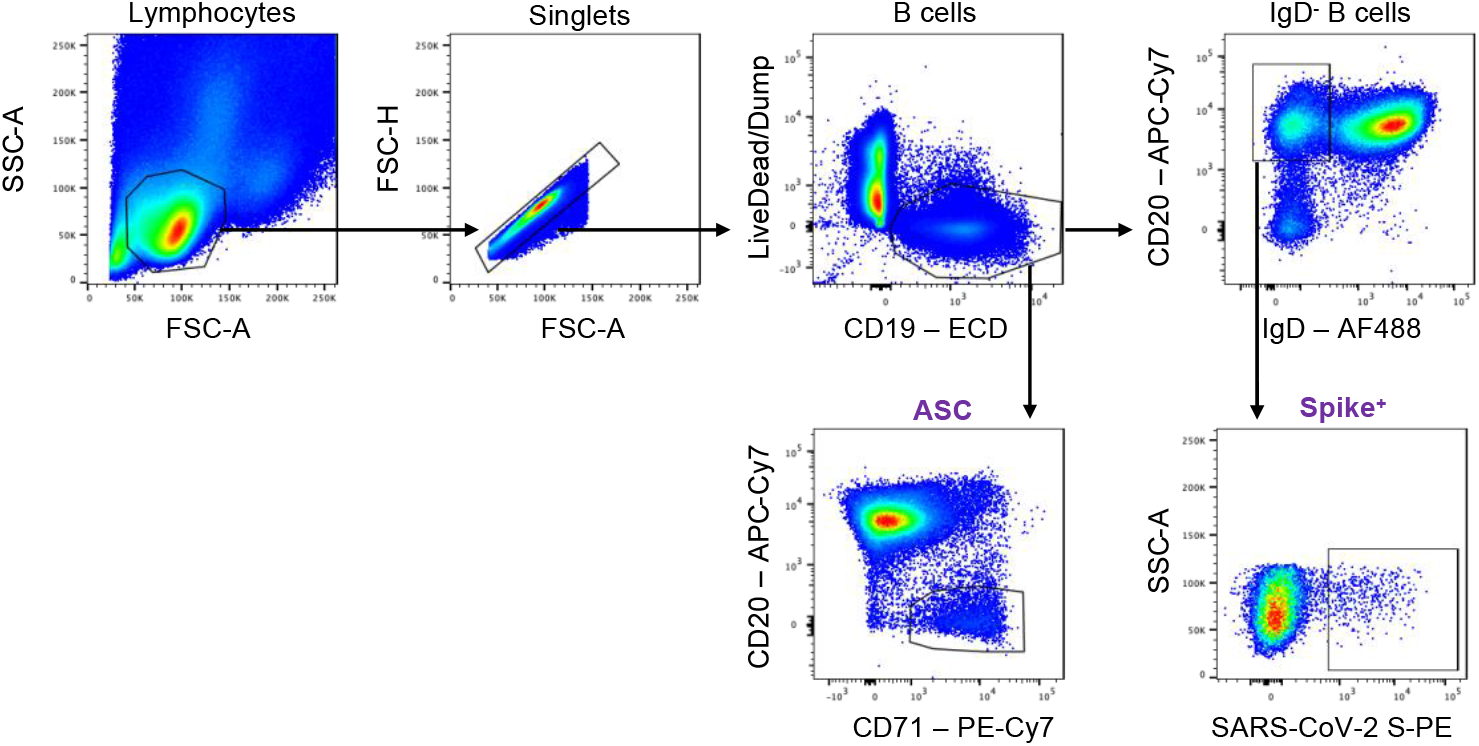
Gating strategy for the analysis of ASCs and S-specific B cells. Lymphocytes were identified by FSC-A vs SSC-A gating, followed by doublet exclusion (FSC-A vs FSC-H), and gating on live CD19^+^ B cells. Antibody-secreting cells (ASCs) were identified as CD20^lo^CD71^+^ within total B cells. Class-switched B cells were identified as IgD^-^CD20^+^ and binding to SARS-CoV-2 S was assessed.

**Supplementary Figure 2.**
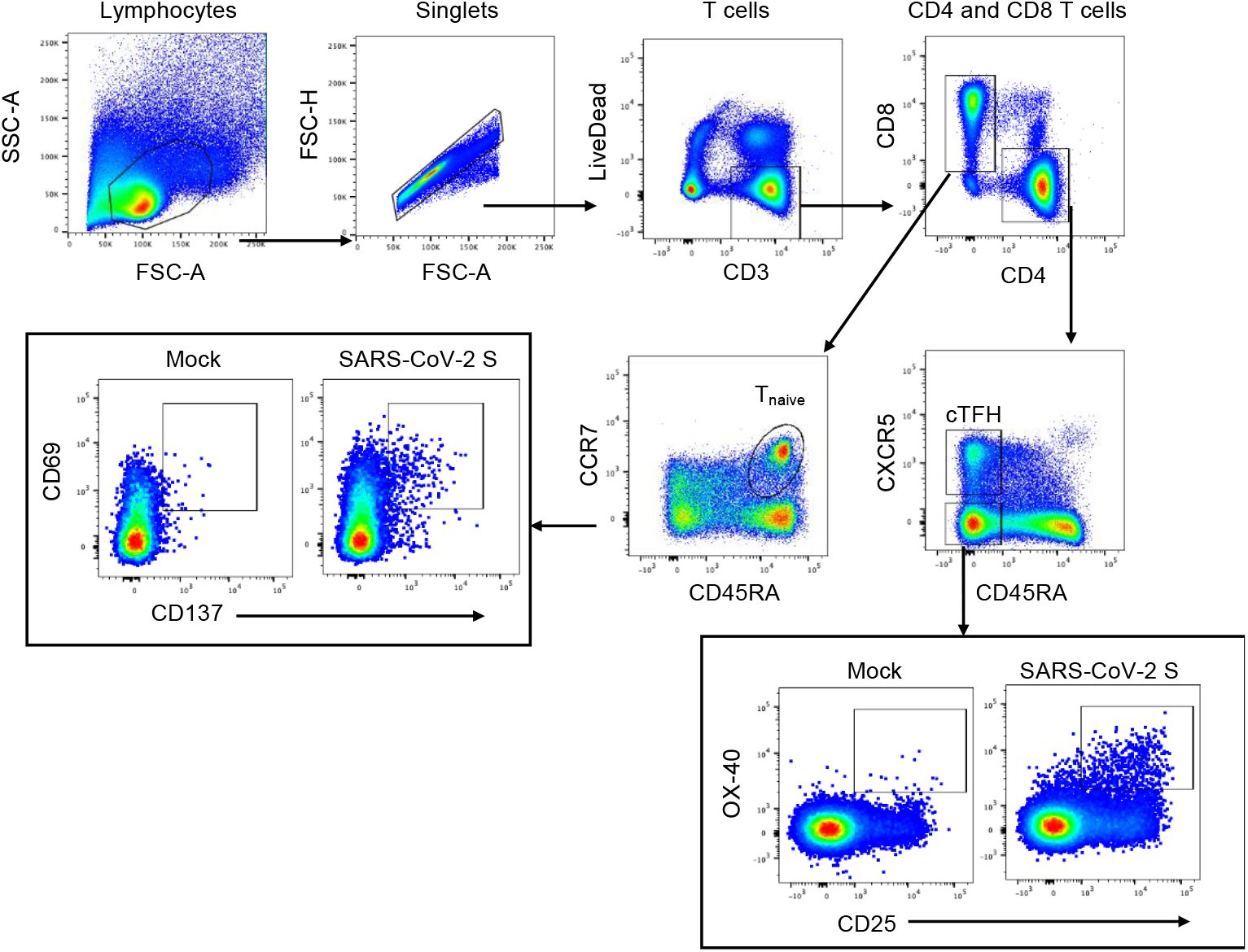
Gating strategy for the analysis of S-specific T cells by AIM. Lymphocytes were identified by FSC-A vs SSC-A gating, followed by doublet exclusion (FSC-A vs FSC-H), and gating on live CD3^+^ T cells, which were further gated as CD4^-^CD8^+^ or CD4^+^CD8^-^ cells. CD4^+^CD8^-^ cells were further defined as cTFH (CXCR5+CD45RA-) or Tmem (CXCR5-CD45RA-), while CD4^-^CD8^+^ cells were further defined as Tmem (non-CD45RA^+^CCR7^+^). Representative FACS plots are shown after stimulation with 5 μg/ml BSA (negative control) or SARS-CoV-2 S protein.

**Supplementary Figure 3.**
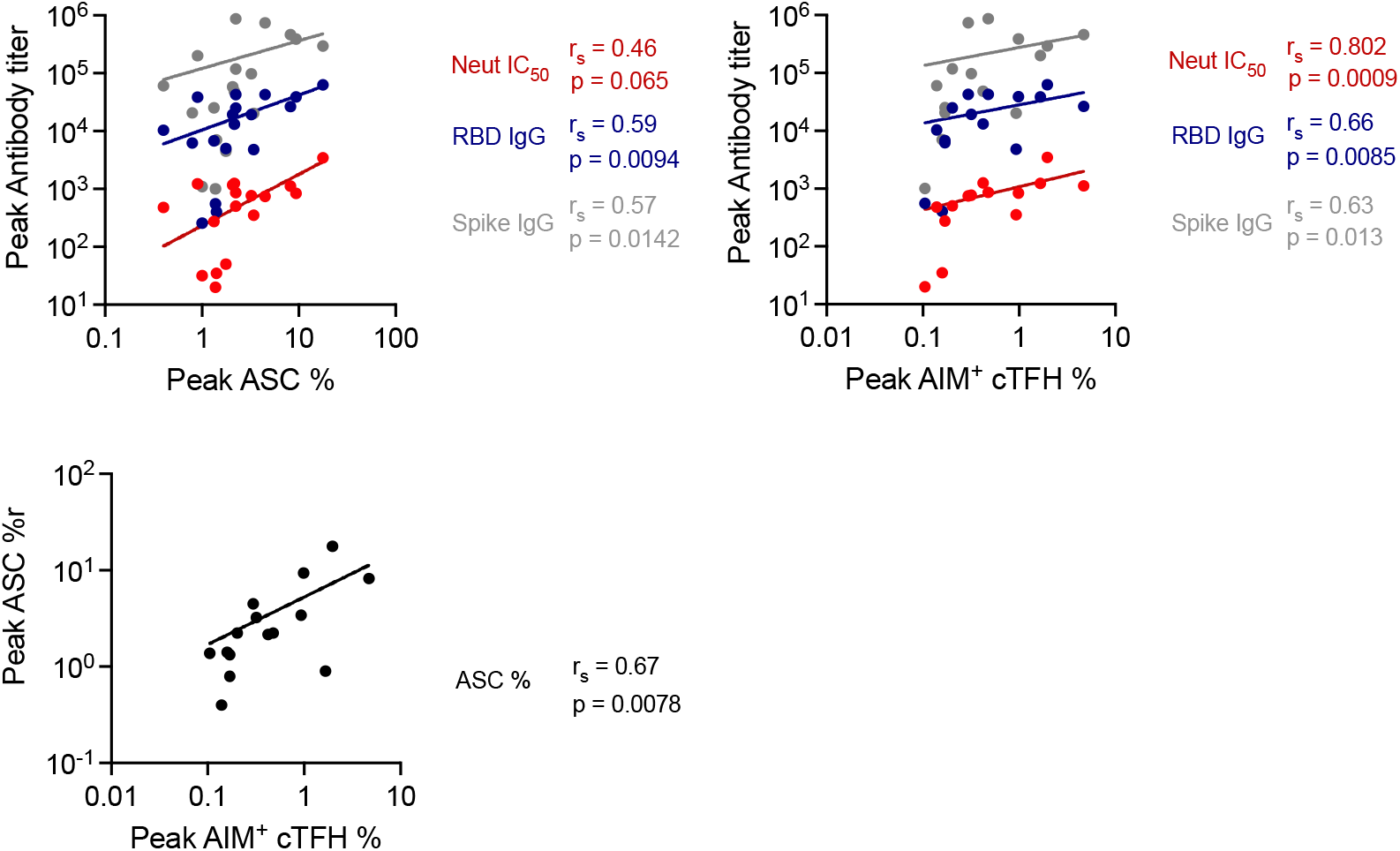
Correlations between cellular and humoral immune responses. Correlation analysis between the peak antibody titres and the peak ASC (n=17 donors) (A) or peak AIM^+^ cTFH cell responses (n=14 subjects). (C) Correlation between peak ASC and peak AIM^+^ cTFH cell responses (n=15 subjects). Spearman correlation coefficients (r_s_) and p values are indicated on the figure.

**Supplementary Figure 4.**
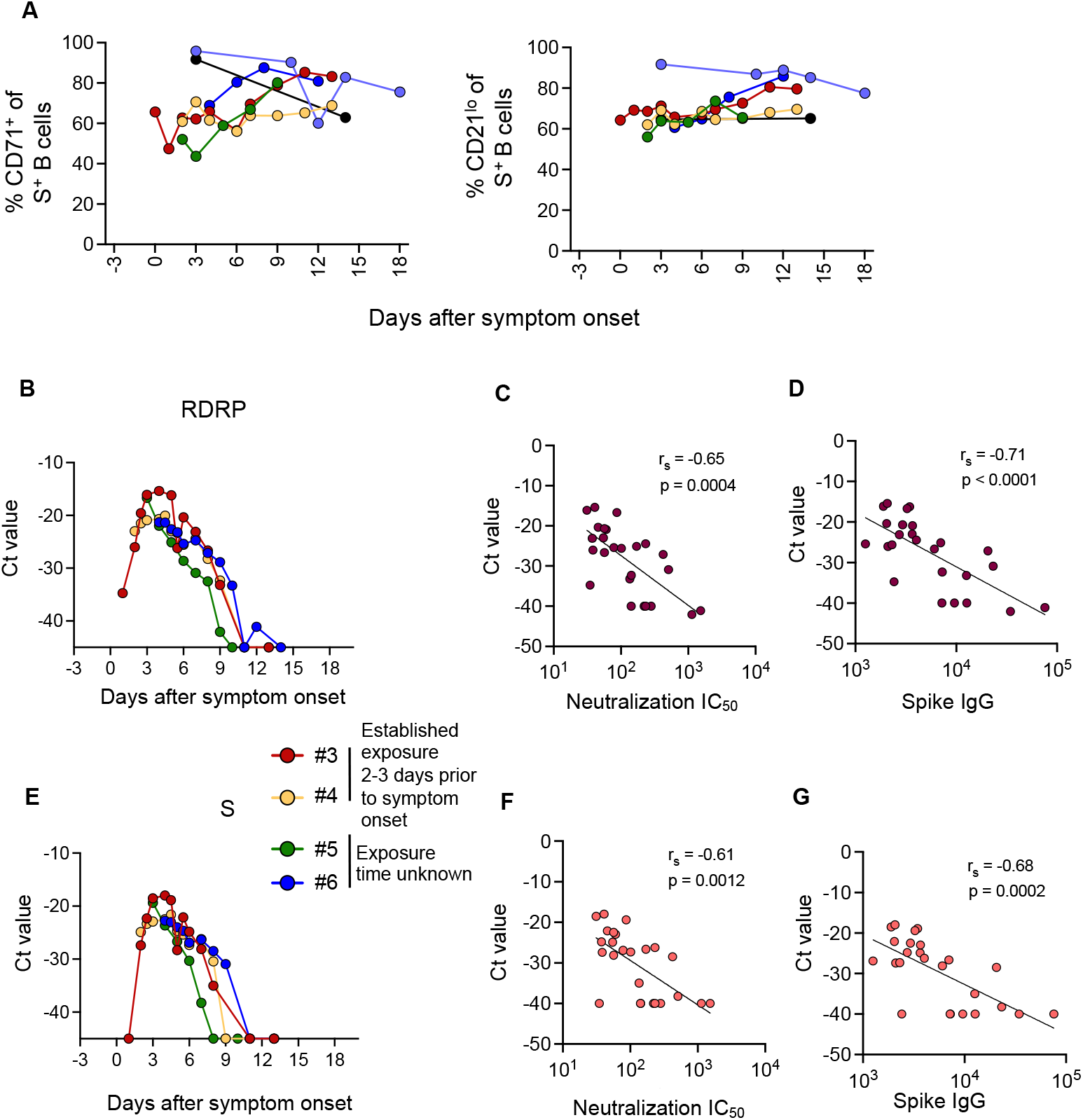
Kinetics of S-specific B cell phenotype and viral load in breakthrough infections. (A) Frequency of activation markers (CD21, CD71) within S-specific class-switched B cells (IgD^-^CD19^+^) in SARS-CoV-2 breakthrough infections (n=6 participants). The x-axis is set as days post symptom onset with negative values indicating the confirmed exposure time for 4 participants (2-3 days prior to onset). (B) Ct values for SARS-CoV-2 RDRP gene in serial nasopharyngeal swabs. (C-D) Correlations between RDRP Ct values and levels of neutralizing antibody titres (C) or Spike IgG antibodies (D). (E) Ct values for SARS-CoV-2 S gene in serial nasopharyngeal swabs. (F-G) Correlations between S Ct values and levels of neutralizing antibody titres (F) or Spike IgG antibodies (G). Spearman correlation coefficients (r_s_) and p values are indicated on the figure, n=25 paired samples from 4 subjects.

**Supplementary Table 1.** Vaccine cohort.

|  | **Seropositive Cohort (n=25)** | **Naïve Cohort (n=8)** |
| --- | --- | --- |
| **Age (Median, IQR)** | 60 (55, 63) | 34 (29, 41) |
| **Gender – Female (n, %)** | 11 (44%) | 4 (50%) |
| **Vaccination, days post-symptom onset (Median, IQR)** | 395 (377, 409) | - |

**Supplementary Table 2.** Estimated parameters of recall following vaccination of seropositive individuals.

| **Vaccine boost** | Estimated parameter (95% CI) |  |  |  |  |  |  |  |
| --- | --- | --- | --- | --- | --- | --- | --- | --- |
|  | Neutralization IC50 | Spike IgG | Spike MBC | ASC | Tetramer | cTFH | CD4 | CD8 |
| activation time (days post vaccination) | 4.85 (4.24 - 5.56) | 5.37 (5 - 5.75) | 6.49 (6.05 - 6.96) | 2.41 (1.27 - 4.58) | 3.53 (2.86 - 4.35) | 3.82 (2.83 - 5.15) | 4.14 (2.1 - 8.14) | 4.13 (2.2 - 7.8) |
| doubling time (days) | 0.74 (0.54 - 1.03) | 0.4 (0.34 - 0.48) | 0.64 (0.38 - 1.07) | 2.06 (1.31 - 3.24) | 0.82 (0.56 - 1.19) | 0.7 (0.32 - 1.56) | 3.43 (1.34 - 8.8) | 0.5 (0.14 - 1.77) |
| fold change (from baseline) | 25.8 (16.6 - 40.2) | 73.8 (40.1 - 136.1) | 2.6 (1.7 - 4.1) | 4.8 (3.1 - 7.6) | 9.7 (4.8 - 28.1) | 11.5 (4.8 - 28.1) | 3.6 (2 - 6.4) | 4.3 (2.1 - 8.7) |

**Supplementary Table 3.** Cohort of SARS-CoV-2 breakthrough infections.

| Donor ID | Age range | Gender | Vaccine | Severity (self-reported) | Time between last dose of vaccination and infection (months) | Days between exposure and symptom onset | Time points analysed for serology (post onset) | Time points analysed for cellular assays | Timepoints analysed for viral load |
| --- | --- | --- | --- | --- | --- | --- | --- | --- | --- |
| 1 | 26-30 | F | ChAdOx1 nCoV-19 | Mild/moderate | 1 | 3 | 3, 14 | 3, 14 | n.d. |
| 2 | 26-30 | M | ChAdOx1 nCoV-19 | Moderate/severe | 1 | 2 | 3, 10, 12, 15, 18, 39 | 3, 10, 12, 15, 18 | n.d. |
| 3 | 26-30 | F | BNT162b2 | Mild/moderate | 4 | 3 | 0, 1, 2, 3, 4, 6, 7, 9, 11, 13, 30 | 0, 1, 2, 3, 4, 6, 7, 9, 11, 13 | 1, 2, 2.5, 3, 4, 5, 5.5, 6, 7, 8, 9, 11, 13 |
| 4 | 26-30 | F | BNT162b2 | Mild/moderate | 4 | 3 | 2, 3, 4, 6, 7, 9, 11, 13, 30 | 2, 3, 4, 6, 7, 9, 11, 13 | 2, 2.5, 3, 4, 4.5, 5, 5.5, 6, 7, 8, 9, 11, 13 |
| 5 | 26-30 | M | BNT162b2 | Mild/moderate | 4 | n/a | 2, 3, 5, 7, 9, 26 | 2, 3, 5, 7, 9 | 3, 4, 5, 6, 7, 8, 9, 10 |
| 6 | 56-60 | F | ChAdOx1 nCoV-19 | Mild/moderate | 5 | n/a | 6, 8, 12 | 3, 6, 8, 12 | 4, 4.5, 5, 5.5, 6, 7, 8, 9, 10, 11, 12, 14 |

**Supplementary Table 4.** Estimated parameters of recall following breakthrough infections.

| Breakthrough boost | Estimated parameter (95% CI) |  |  |
| --- | --- | --- | --- |
|  | Neutralization IC50 | Spike IgG | Spike MBC |
| Activation time (days post exposure) | 7.22 (7 – 8.91) | 8.21 (6.9 – 9.8) | 9.3 (7 – 13) |
| doubling time | 1.6 (1.3 – 2) | 1.1 (0.68 – 1.74) | 3.17 (2.2 – 4.6) |
| fold change (from baseline) | 12.2 (5.9 – 25.2) | 15.8 (3.9 – 64.4) | 3.92 (1.6 – 9.5) |

